# A qualitative study exploring the impact of the COVID-19 pandemic on People Who Inject Drugs (PWID) and drug service provision in the UK: PWID and service provider perspectives

**DOI:** 10.1101/2022.01.24.22269530

**Authors:** Tom May, Jo Dawes, Daisy Fancourt, Alexandra Burton

## Abstract

**Background:** People Who Inject Drugs (PWID) are subject to distinct socio-structural inequalities that can expose them to high risks of COVID-19 transmission and related health and social complications. In response to COVID-19 mitigation strategies, these vulnerabilities are being experienced in the context of adapted drug treatment service provision, including reduced in-person support and increased regulatory flexibility in opioid substitution therapy (OST) guidelines. This study aimed to explore the longer-term impact of the pandemic on the health and wellbeing of PWID in the UK, including provider and client experiences of treatment changes.

**Methods:** Interviews were conducted with 19 PWID and 17 drug treatment providers between May – September 2021, recruited from third-sector drug services in the UK. Data were analysed using reflexive thematic analysis.

**Results:** Most participants expressed ongoing fears of COVID-19 transmission, although socio-structural inequalities limited the contexts in which physical distancing could be practised. In addition, virus mitigation strategies altered the risk environment for PWID, resulting in ongoing physical (e.g. changing drug use patterns, including transitions to crack cocaine, benzodiazepine and pregabalin use) and socio-economic harms (e.g. limited opportunities for sex work engagement and income generation). Finally, whilst clients reported some favourable experiences from service adaptations prompted by COVID-19, including increased regulatory flexibility in OST guidelines, there was continued scepticism and caution among providers toward sustaining any treatment changes beyond the pandemic period.

**Conclusions:** Whilst our findings emphasize the importance of accessible harm reduction measures attending to changing indices of drug-related harm during this period, there is a need for additional structural supports to ensure pre-existing disparities and harms impacting PWID are not exacerbated further by the conditions of the pandemic. In addition, any sustained policy and service delivery adaptations prompted by COVID-19 will require further attention if they are to be acceptable to both service users and providers.

## Introduction

In response to the COVID-19 pandemic, governments worldwide implemented various mitigation measures - including physical distancing, mobility constraints and the closure of social, business and educational settings - in efforts to suppress the virus. Whilst these measures resulted in significant disruption to the lives of many (Brooks et al., 2020; Pierce et al., 2020), their effects varied across populations and disproportionally impacted some of the most marginalised members of society (Bambra & Lynch, 2021; Bambra, Riordan, Ford, & Matthews, 2020). This included People Who Inject Drugs (PWID), a population subject to pre-existing socio-structural inequalities (e.g. economic disadvantage, housing instability, stigma) that exposed them to high risks of COVID-19 transmission and related health and social complications (Bennett, Townsend, & Elliott, 2021; Kesten et al., 2021; Vasylyeva, Smyrnov, Strathdee, & Friedman, 2020).

Emerging research has documented the initial health and social impacts of the pandemic on PWID. Firstly, whilst mitigation measures were implemented in efforts to minimise contact and transmission, they disrupted daily routines and access to health and social care, and increased adverse mental health impacts, including isolation, boredom and anxiety (Bennett et al., 2021; Kesten et al., 2021; Roe et al., 2021). Further, travel restrictions and border closures resulted in unstable drug markets and variable drug supplies, with some evidence of poly-drug use and substance substitution in response to temporary shortages at local levels (Croxford et al., 2021; Morin, Acharya, Eibl, & Marsh, 2021). Finally, widespread business closures and a decline in opportunities for informal income-generating activities (e.g. begging, theft, sex work) exacerbated material and economic hardship for PWID, increasing vulnerability to both drug (e.g. opioid withdrawal) and health (e.g. hunger) related harms (Bennett et al., 2021). A combination of these factors may be behind increased risk behaviours (e.g. syringe sharing, poly-drug use) (Nguyen & Buxton, 2021; Perri et al., 2021) and the acceleration of fatal and non-fatal opioid overdoses since the introduction of social distancing measures worldwide (Friedman & Akre, 2021; Glober et al., 2020; Rodda, West, & LeSaint, 2020; Slavova, Rock, Bush, Quesinberry, & Walsh, 2020).

In response to social distancing measures implemented during the earlier stages of the pandemic, many services reconfigured treatment and support for PWID to limit daily clinical encounters and reduce chances of virus transmission. For example, greater regulatory flexibility in opioid substitution therapy (OST) guidelines, including a shift from the daily supervised consumption of agonist medications (e.g. methadone and buprenorphine) to the provision of risk-assessed ‘take-home doses’, was introduced in the UK and US (Department of Health and Social Care, 2021; SAMHSA, 2020)^1^. Further measures - including the home delivery of harm reduction equipment (e.g. naloxone, injecting equipment), mobile outreach and expanded telephonic and telehealth services - were also established in attempts to increase treatment access in the context of reduced service availability (Aronowitz et al., 2021; Courser & Raffle, 2021; Mehtani et al., 2021; Nordeck, Buresh, Krawczyk, Fingerhood, & Agus, 2021). Given how treatment engagement and retention is often compromised by inaccessibility and – in the case of OST - the daily burden of supervised consumption (Frank, 2021; Frank et al., 2021; Hall, Le, Majmudar, & Mihalopoulos, 2021), the pandemic presents a context in which these particular issues could be addressed and improved, at least temporarily. Although service providers reported concerns that adaptations may have decreased quality of care and led to increased instances of medication diversion and overdose (Hunter, Dopp, Ober, & Uscher-Pines, 2021), emerging evidence from survey data generally conveys favourable outcomes among clients with limited occurrences of medication diversion or misuse (Figgatt, Salazar, Day, Vincent, & Dasgupta, 2021; Joseph, Torres-Lockhart, Stein, Mund, & Nahvi, 2021). Such contradictions reflect pre-existing debates on the use of telehealth modalities in healthcare (Scott Kruse et al., 2018), as well as provider/client discussions regarding the optimal delivery of OST (Anthony et al., 2012; Frank, 2021).

Given how social and economic upheavals prompted by COVID-19 disproportionately impact the health and wellbeing of the most vulnerable (British Academy, 2021), the effects of the pandemic on PWID are likely to be long-lasting and are now only beginning to emerge. In this context, there remains a need for research with PWID and service providers beyond the initial stages of the pandemic, including any lasting and sustained impacts on drug-use patterns, drug-related harms and mental health previously identified (Bennett et al., 2021; Kesten et al., 2021). Of further interest is how both providers and clients are continuing to respond to COVID-initiated service adaptions, including the continuation/discontinuation of remote working practices (e.g. telehealth, reduced in-person appointments) and the relaxation of OST regulations. Whilst studies have reported early insights from clients toward some of these changes (e.g. Kesten et al., 2021), the perspectives and reactions of providers to altered treatment delivery are mainly absent, particularly in the UK. Understanding how service providers responded to the pandemic - including their comfort and willingness to use new forms of service delivery - can provide important practice and policy insights for whether seemingly temporary service responses to COVID-19 persist beyond this period.

Therefore, the current study investigates the longer-term impacts of the pandemic on the health and wellbeing (including drug-related harms and risk behaviours) and everyday lives of PWID, as well as their experiences of treatment changes from the perspectives of both PWID and service providers.

## Methods

The research employed a qualitative design using semi-structured interviews with PWID and service providers from third-sector drug services in England and Scotland. The study formed part of the UCL COVID-19 Social Study (UCL, 2021), which explores the psychosocial effects of COVID-19 and associated restrictions on people living in the UK. Participants were interviewed about their experiences throughout the pandemic, including any implications for substance use, treatment engagement and delivery, and mental health and wellbeing. Ethical approval was provided by University College London research ethics committee [Project ID 6357/002].

The study was conducted between May – September 2021, a period when COVID-19 measures in England and Scotland were beginning to ease. COVID-19 measures were relaxed in England and most areas of Scotland on May 17^th^ 2021, allowing meetings of up to six people indoors and the majority of the indoor economy to reopen. The easing of these measures followed a national lockdown period between January and April 2021 that curbed social mixing outdoors and closed non-essential retail. The bulk of legal COVID-19 restrictions were lifted in England and most areas in Scotland on 19^th^ July 2021, although other types of restriction, notably self-isolation and international quarantine restrictions, remained. Appendix 1 provides a summary of the lockdown measures during the period of fieldwork.

### Sample and recruitment

Convenience sampling was used to recruit both groups of participants, although as more interviews were conducted, we specifically advertised for under-represented characteristics within our sample to ensure diversity of gender, age, ethnicity and occupation in the case of service providers. In practice, this involved recruiting participants through several methods, including via social media and the UCL COVID-19 Social Study (including its newsletter and website). A national (with services across the UK) and two local third-sector drug treatment services (located in Bristol and London) also advertised the research via bespoke posters and fliers within service settings and helped promote the research to eligible participants on our behalf. Clients were eligible to participate if they were (i) a current injecting opioid user or had been injecting opioids at some stage during the pandemic, (ii) aged over 18, and (iii) living in the UK. Service providers could participate if they (i) worked directly with PWID in drug services, (ii) were aged over 18, and (iii) living in the UK.

Members of the research team provided eligible participants with details about the purpose of the research in writing and verbally and informed them that their involvement was voluntary. In addition, all participants signed a consent form, and demographic details (e.g. age, gender, ethnicity) were obtained. This information was received via email for participants who were interviewed remotely, or ‘in person’ if interviewed face-to-face.

### Data Collection

Interviews were conducted by TM (research fellow in social science), and JD (physiotherapist and research fellow in health inequalities) via telephone/video call (n=21) or in-person (n=15). All interviews were conducted individually apart from one interview, which was conducted with two PWID who requested to be interviewed together. Interviews followed a semi-structured topic guide for each participant group, which enabled data collection on the impact of the pandemic on substance use, treatment engagement and delivery and mental health and wellbeing (see Appendix 2 and 3 for full topic guides). Interviews lasted an average of 38 minutes, were digitally recorded and transcribed verbatim by a professional transcription service. All interviews with service providers were conducted remotely (n=17) while the majority of client interviews were conducted face-to-face within either a drug service (n=9) or hostel facility where drug service clients resided (n=6). These interviews took place in a ventilated room and the researcher followed social distancing guidelines. The remaining client interviews were conducted remotely (n=4). Monetary compensation in the form of a £10 high street or supermarket voucher was offered to thank participants for their involvement. Data collection was conducted alongside data analysis and continued until theoretical saturation occurred across the entire sample (i.e. the point at which data emerged consistently or where no further data would develop new properties, categories or findings (Fusch & Ness, 2015)).

### Data analysis

Transcripts were uploaded to NVivo version 12 software after de-identification. We used reflexive thematic analysis (Braun & Clarke, 2019, 2021) to analyse the data. This began with TM and JD independently reading and coding the same three transcripts. They then met to discuss and compare topics of potential significance to the research objectives. Following this process, TM then read and coded the remainder of the dataset, identifying further aspects of data that were then developed into a set of initial codes based on content grounded in the participant quotes. These codes were subsequently used to generate a set of key themes after being reviewed and analysed, which resulted in some codes being combined to form their own themes or sub-themes. To assist with this process, the research team met weekly as part of an ongoing iterative process to refine any new codes or themes produced. The use of two sources of qualitative data allowed for the triangulation and contextualisation of service user accounts with service provider interviews.

## Results

### Participant characteristics

We interviewed 36 participants (19 clients and 17 service providers). All client participants were recruited through third-sector sector drug services; four were recruited through a service based in London and 15 through a service in Bristol. The average age of clients was 40 (range 24-59), with just over half identifying as female (n= 10). Most clients were White British (n=13) and at the time of interview most were temporarily housed, either in a hostel or with friends/family. All clients reported current or recent use of heroin or heroin and crack cocaine, and instances of alcohol, benzodiazepine and pregabalin use were also common. Table 1 provides an overview of client characteristics.

**Table 1.** Characteristics of PWID.

| **Demographics** | **Range/n (mean)** |
| --- | --- |
| **Age** | 24-59 years (40.1 years) |
| **Gender** |  |
| Female | 10 |
| Male | 9 |
| **Ethnicity** |  |
| White British | 13 |
| Black or Black British Caribbean | 2 |
| White and Black Caribbean | 2 |
| White Other | 2 |
| **Housing Situation** |  |
| Hostel | 8 |
| Rented (council) | 6 |
| Rented (private) | 2 |
| Temporary with friend/family | 2 |
| Street Homeless | 1 |
| **Substance Use\*** |  |
| Heroin and crack cocaine | 18 |
| Diazepam | 4 |
| Alcohol | 4 |
| Pregabalin | 4 |
| Spice | 2 |
| Heroin | 1 |

**\*Refers to use of all reported substances**

Seventeen interviews were conducted with service providers from a single third-sector drug treatment provider with services located across England and Scotland. Service providers were from various occupational backgrounds within the drug and alcohol field, including clinical, management and frontline staff. The average age of service providers was 45 (range 28-67), with just over half identifying as male (n=9). The majority were White British (n=13). Table 2 provides an overview of service provider characteristics.

**Table 2.** Characteristics of service providers.

| **Demographics** | **Range/n (mean)** |
| --- | --- |
| **Age** | 28-67 years (45.5) |
| **Gender** |  |
| Male | 9 |
| Female | 8 |
| **Ethnicity** |  |
| White British | 13 |
| Black or Black British African | 2 |
| White Other | 2 |
| **Occupation** |  |
| Drug Service Team Leader/Services Manager | 7 |
| Drug Service Worker (Recovery Coordinator) | 5 |
| GP with Addiction Specialism | 2 |
| Substance Use Nurse | 2 |
| Consultant Psychiatrist | 1 |
| **Location** |  |
| London and South East | 6 |
| North West | 4 |
| East Midlands | 2 |
| Yorkshire and Humber | 2 |
| Scotland | 1 |
| South West | 1 |
| West Midlands | 1 |
| **Years of experience** |  |
| 1-5 years | 2 |
| 6-10 years | 8 |
| 10 years + | 7 |

Three primary themes were identified: (1) ongoing fears of COVID-19 infection but limited possibilities for guideline adherence; (2) increased social and drug-related harms, and; (3) experiences of service adaptations. Themes are shown in Figure 1, along with their respective subthemes.

**Figure 1.**
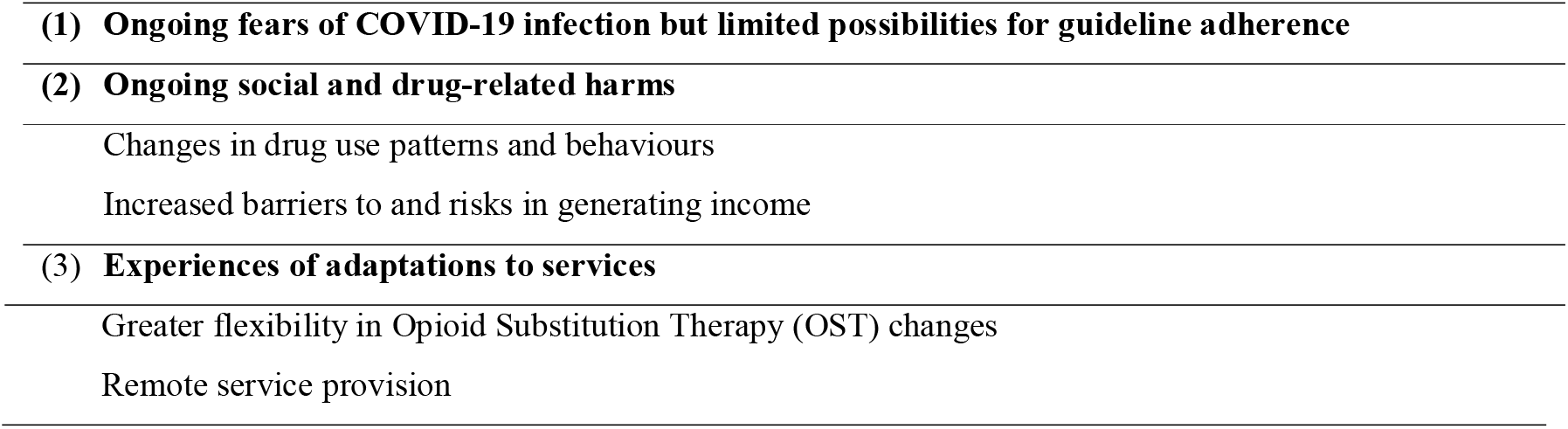
Key themes.

### Ongoing fears of COVID-19 infection but limited possibilities for guidelines adherence

Most clients described how their adherence to social distancing had declined since earlier in the pandemic (*like at first most people did take it seriously, [but] on the second one [lockdown], the drinkers around my way didn’t take it seriously*, client 2, F, 46-50). This was often linked to feeling less worried about the consequences of COVID-19 due to previous possible infection (*I reckon us streeties have probably had it and had it mutate in us so many times*, client 2, F, 55-60) or knowing of no infection among others (*I’m not really that fussed. I haven’t seen anyone with it, I don’t know anyone who’s had it*, client 6, M(ale), 31-35) There was agreement among service providers that infections had been lower than expected among clients, which was both surprising and unexpected given earlier concerns:

> *I guess that was something that was a relief in a way but also a bit of a surprise that they weren’t as severely affected as we felt they might be…I think we felt the risk of more severe consequences would have been higher for more people but we were pleasantly surprised I guess that they weren’t* (consultant psychiatrist, M, 36-40)

Most participants, however, attributed declines in adherence to daily necessities that were unconducive to sustained periods of distancing. This included how more immediate everyday concerns – including avoiding withdrawal and income generation – were often prioritised over compliance with measures. Those involved in street-based sex work, for example, reported ceasing activity at first but later returning, despite concerns about contracting COVID-19 (*I did feel scared. I stopped sex working in the first wave…I got really scared that I was actually going to die, because I knew I probably would if I got it*, client 3, F, 21-25). Although worried, this participant saw continued engagement in sex work as necessary in the absence of sufficient social and economic support:

> *Yes, it put not just me but a lot in dire straits really. Because the thing is our income was cut literally overnight. There was no warning. And then there was no furlough scheme for sex workers. Benefits went up £20 a week. But what did that do, nothing. It wasn’t a dent* (client 3, F, 21-25)

COVID-19 fears also remained and were described within the context of accommodation settings. At the time of fieldwork, most clients were housed temporarily in hostels. Both clients and providers noted how, despite COVID-19 measures being in place, adherence was now variable (*When you walked in, they would be like, “Hand sanitiser when you come in. Mask, fresh mask when you come in,” but no one would really stick to it*, client 5, M, 26-30). Older clients and those with pre-existing health conditions reported feeling vulnerable to infection within accommodation settings, particularly as communal spaces were often ‘overcrowded’ and ‘cramped’. This often elevated fears of transmission:

> *We’re in a small block of flats…we know there is no way that if the disease got in the block that it wouldn’t get caught by us. It’s got communal lifts, communal stairways, communal washing machines, so we just prayed to God that it didn’t get everywhere* (client 4, M, 46-50)

### Ongoing social and drug-related harms

Clients and service providers reported how earlier lockdown measures had exacerbated several indices of drug-related harms, which were found to have ongoing and sustained impacts on drug use patterns and behaviours, and the ability to generate income.

### Changes in drug use patterns and behaviours

Both providers and clients agreed that lockdown measures had made it more difficult for clients to source heroin. Obstacles to conducting drug transactions in public spaces were described, as were reports of local drug supply shortages and increased adulteration. Although some clients responded to these constraints by ceasing or reducing their heroin use (*I’m not doing that recently because of how crap it is, how small it is…there’s just no point*, client 8, M, 46-50), some reported sourcing other substances in attempts to alleviate withdrawal symptoms:

> *I started buying Fentanyl patches online. And then I started buying pregabs*…*so, I started taking them, just to keep me going so I wasn’t withdrawing* (client 3, F, 21-25)

Some participants indicated that drug market volatility had ended and purity re-established by the time of interview, and that they had therefore returned to the sole use of heroin. Some reported how concerns about the harms associated with previous poly-drug use contributed to this pattern:

> *I was then drinking, doing benzos…and [keyworker] was like, ‘Jesus Christ, you’ve got to stop something, otherwise you’ll end up overdosing by accident’. So, I did. I stopped the alcohol and benzos, because they weren’t really my primary substances I used to use* (client 3, F, 21-25).

Others, though, reported how changes in drug-use patterns were being sustained beyond earlier periods of the pandemic. Some explained how benzodiazepines and pregabalin were ‘*just so addictive*’ (client 4, M, 46-50) and were unable to cease using them (*At the moment I’m trying to stay away from it [heroin]. But to be honest we’ve been using Pregabs…they’re hard to come off… I’ve been taking a whole strip every day for a while, I can get them every day*, client 8, M, 46-50). Although common before the pandemic, there was agreement in service provider narratives that the use of benzodiazepines and pregabalin were now more widespread because of earlier substitution:

> *though self-referring for drug issues has pretty much remained, there’s not been too much difference. The types of drugs people are using has changed…more of the street drugs, more of the street Benzo’s (Recovery Coordinator, F, 51-55)*

Client and service provider narratives highlighted the consequences of these transitions, including the increased potential for drug-related harms. Whilst no reports of overdose were stated, anecdotal reports of increased drug-related deaths during this period were often linked to the use of illicit benzodiazepines (*There’s a huge street Benzo problem in [location], and it’s one of the major factors driving the drugs deaths rate. They’re very easily available and very cheap*, recovery coordinator, F, 51-55). Some clients also described how the use of benzodiazepines and pregabalin was now more problematic than previous heroin use, with reports of blackouts, memory loss and tolerance issues:

> *The benzos and the prescription drugs, that’s been a problem because of the blackout, it terrifies me to be honest…I don’t remember things being quite as bad as that when I was injecting gear, and like I have had black outs, OD’d a few times, but not like not accounting for hours of time, waking up in different places…I would feel huge anxiety and I have got into a bit of a problem* (client 9, F, 41-45)

Additionally, some clients who would normally use only heroin noted how they had developed continuing patterns of crack cocaine use since the onset of the pandemic. Whilst providers suggested that increased availability and recent fluctuations in local supply may have facilitated this trend (*Within the last two months I’ve noticed more and more people coming to me with crack-cocaine problems. So there’s been a flood of that, I’m not sure what’s behind that, but there has been a flood of crack in [location] specifically*, recovery coordinator, F, 51-55), clients explained how the short-term and intense effects of crack cocaine encouraged the compulsive use of the substance. Some client narratives suggested a development of rapid increases in tolerance and the need to now use more frequently:

> *[When] you have a bit of crack, it feels quite good at first but it never keeps its promises up. And the crapper the crack is the quicker it goes – like you can do a pipe and literally be doing one straight after* (client 11, M, 41-45)

### Increased barriers to and risks in generating income

A small number of clients reported job losses earlier in the pandemic, and subsequent ongoing difficulties reengaging with formal or casual employment due to a lack of opportunities. Income constraints meant some were now reliant on Universal Credit, which represented a substantial decrease in income compared to before the pandemic. The daily economic demands of drug use therefore became more burdensome, with problems reported in relation to obtaining funds for travel, self-care and drug acquisition:

> *So, I just had Universal Credit. And because I was under 25, it was £250 a month. It’s fucking inhumane. I don’t know what people expected me to do. I couldn’t afford transport to my appointments. I couldn’t then eat. And then obviously fucking drugs on top of that, it was ridiculous* (client 3, F, 21-25)

Most clients were reliant on informal income-generating activities to meet basic subsistence needs, including shoplifting, sex work and begging. Some described how these activities had been disrupted by earlier lockdown measures, impacting their ability to generate income later in the pandemic. For instance, earlier shop closures and heightened security made shoplifting harder and increased the chances of detection. There were some isolated accounts of clients being caught during these earlier periods, which later restricted their ability to return to stores once relied upon for shoplifting (*we can’t go near none of them…It just absolutely destroyed any shoplifting*, client 12, M, 36-40). Others reported how opportunities for begging were now limited compared to before the pandemic because “*people always say they don’t carry cash any more*” (client 13, F, 31-35). Women engaged in sex work also described a reduced demand for services once social distancing restrictions were eased, which was linked to customer health concerns (*because obviously the fucking pandemic, they were scared of us*, client 3, F, 21-25) and an accelerated shift toward online sexual services in recent months (*a lot of people [clients] have gone on the internet now anyway*, client 7, F, 41-45).

Income constraints compelled some to generate funds through high-risk methods to alleviate withdrawal symptoms. In haste, some displaced shoplifting to stores they usually avoided, which increased the risk of arrest. For instance, one participant resorted to shoplifting at a local store where her presence was known and more visible, which resulted in her being caught:

> *I never had any earning power anymore…I got caught in <Supermarket> and I don’t normally shit on my own doorstep…I got caught because that was very recent and I don’t normally shoplift in <Supermarket>. But I think it was a day when I was using with somebody else, it wasn’t my money. So, it started the ball rolling and I thought fuck, I need some earning. So, I just ran in there quickly* (client 2, F, 56-60)

Some women also described entering into relationships with men for financial support. For instance, whilst one participant described herself as “*usually quite fiercely independent. Like, no, I make my own money, I buy my own drugs*” decreases in income meant “*I started relying on other people for drugs and income. I started relying on blokes. They’d give me drugs, or they’d get money for drugs*”. This led to an exploitative relationship whereby her partner exercised control over the resources she generated through sex work:

> *And then I’d gravitate towards them [men], because I needed drugs. And I’d never done this before…and I think that’s when it got a bit exploitative, because they then started demanding things in return. And then that’s when it got out of hand. So, I’ve never been in that situation really before…*.*And then I started working for him for drugs, which I don’t think would have happened otherwise* (client 3, F, 21-25)

A reduced demand for street-based sex work once social distancing restrictions eased meant those continuing with or returning to street-based sex work resorted to lowering prices (*you were having punters wanting to give you £10 and all that shit*, client 2, F, 56-60; *there were girls out there doing it for £10* (client 7, F, 41-45). A reduced ability to assert a minimum price meant some were not only at risk of economic exploitation, but the ability to negotiate safe sex practices:

> *They would ask for more, and they’d demand more, or they’d push the boundaries more, because they could, because they knew you needed the money. And no one else was there, so they’d demand things that you didn’t want to do, or try and force you to do it anyway* (client 3, F, 21-25)

### Experiences of Adaptations to Services

Clients and providers described their experiences of service adaptations initiated in response to the pandemic. Although a range of alterations were reported (e.g. home delivery of harm reduction equipment), some of the most significant changes related to the relaxation of OST regulations and shift toward forms of remote provision, including telephone or video calls for psychosocial services and assessments. In line with clinical guidance (Department of Health and Social Care, 2021), most clients who transferred off supervised consumption in the initial phase of the pandemic had returned to daily supervision by the time of interview, and some services had recommenced in-person services. The timing of these changes allows for the exploration of how earlier alterations in services were experienced and how both clients and service providers received the return to previous treatment regimes.

### Greater Flexibility in Opioid Substitution Therapy (OST) Changes

In the initial phase of the pandemic, most services transitioned clients from the daily pick up of OST mediations to either weekly or fortnightly collection. In doing so, most providers reported unease and raised concerns about the loss of benefits linked to the ‘structure’ of supervised consumption, including peer support and regular monitoring, as well as the potential for misuse and diversion:

> *The initial thought of mine was, oh my god, everyone’s going to die, everyone’s going to overdose, everyone’s going to drink it all in one go, they’re going to give it to a friend, they’re going to die, their dog’s going to drink and the dog’s going to die* (team leader 1, F, 25-30)

In response, some service providers conducted risk assessments, a task which helped determine client suitability for less frequent OST collections (*the recovery workers would identify people who they felt were high risk on their caseloads and then we’d discuss them and identify which ones of those were the highest risk and therefore needed to be on daily pickup*, consultant psychiatrist, M, 36-40). Some providers also described enacting further measures to reduce harm, including providing safe storage boxes for methadone, increased naloxone distribution, and the home delivery of OST medications to clients unable to attend pharmacies. This method allowed service providers to also ‘check in’ with clients and provide other supports:

> *Coming to their door…we were able just to spend a bit of time talking to them. If necessary we were able to take food out to them. We were able to take clean needles and the lockbox…if people had children in the home we were taking like children’s activity packs when they were off from school. You know trying to provide a more comprehensive service than just going, “Here’s your prescription, here’s your methadone* (recovery coordinator, F, 51-55)

Whilst reports of serious harm were limited, some instances of misuse were described, including “lost” bottles (*we had a few saying,“Oh I dropped the bottle”*, team leader, F, 51-55) and increased local diversion (*I’ve seen semi-recently in the last year…clients being able to buy illicit Methadone. So, that’s come from there being an excess on the streets*, substance misuse nurse, M, 31-35). However, service providers reported how these occurrences were largely kept under control by reverting clients to previous prescribing regimes if misuse or safety concerns were identified:

> *We gradually worked out who needed let’s say closer supervision for their own safety, right? And when people say, “Why are you putting me in daily supervise? That’s a punishment*.*” I’d say, “No, it’s not a punishment, this is for your own good, this is, I hate to say this, this is to keep you safe* (GP, M, 65-69)

Clients noted how the change to fortnightly pick-ups eliminated negative experiences associated with dispensing environments, including shame (*It’s a bit embarrassing having to check your mouth and that, do you know what I mean? One because I’ve got bad teeth and two I’m nearly 50*, client 4, M, 46-50) and possible theft of medication (*I used to be in that chemist there, and it’s terrible there, and there used to be a gang of them outside either trying to get meds off you or they want to swap for you*, client 4, M, 46-50). Others favoured the more considered and patient-centred approach to treatment, including the increased autonomy they had over dosage and titration. For instance, some reported splitting their methadone doses over the course of the day, which enabled them to manage their drug use more efficiently:

> *Yeah, because then I could limit what I could have. For instance, I was having an 8ml in the morning and then a 2ml five hours later, and then maybe another 2ml, and then another 2ml before I went to bed* (client 5, M, 26-30)

As social distancing measures relaxed, most services reverted to previous OST protocols, including daily supervised pick-ups (Department of Health and Social Care, 2021). The experience therefore offered an opportunity for services and clients to reflect on the future delivery of OST. As the findings above attest, the pandemic highlighted the inflexibility associated with daily supervised consumption for most clients. Some clients who transitioned back to pre-COVID regimens voiced frustration at doing so. For some, it was a nuisance that jeopardised their engagement with treatment:

> *Yes, because I can manage my meds myself. Like some days I don’t go down there because I can’t be bothered to go down there, or I can’t be bothered to queue up, you know…It’s queueing up and it’s all long, long, long, do you know what I mean? Sometimes I just can’t be bothered, can’t face it* (client 4, M, 46-50)

Service providers reported mixed feelings towards the return to daily supervised dosing based on both their and client experiences; of being generally supportive toward longer term reforms to OST prescribing, or favouring a return to the ‘structure’ of daily supervision. For instance, despite initial concerns, some providers became more comfortable with flexible dosing and agreed that discussions regarding future OST changes were warranted, given how only limited incidents of harm were reported. That most clients had ‘proved’ themselves capable of managing take-home doses also meant some service providers were now more trusting of them:

> *There can be this tendency because of the nature of the people’s support needs to be a bit mistrusting of them…the pandemic forced us into a position where we had to be more trusting of people…there’s massive positives to that. So we’ve become more allowing of people to just get on with their lives and may be work a bit more freely with them around whatever’s going on for them and then just be okay with that* (recovery coordinator, M, 31-35)

Nevertheless, many providers still reported concerns surrounding quality of care and client risk and were hesitant to advocate long-term changes. Some also felt that the conditions of the pandemic allowed for more favourable OST outcomes to be achieved and that diversion and misuse would return once social distancing measures had been fully relaxed:

> *I think we need to take those changes in the context of the pandemic when people were mingling a lot less and people were probably keeping themselves to themselves a lot more. So, you know if you went back to pre-pandemic levels of mingling and social interaction and people were having 14 days’ worth of take home methadone, Buprenorphine etc, my feeling is that there would be a lot more diversion in that context* (consultant psychiatrist, M, 36-40)

### Remote service provision

Services also responded to earlier social distancing measures by transitioning to forms of remote provision, including the use of telephone or video calls for psychosocial services and assessments. Although some reported returning to some in-person services by the time of fieldwork, both clients and providers noted several reasons for why they felt elements of remote provision should be continued. For instance, some clients enjoyed the increased accessibility that online sessions enabled, particularly those with mental health issues who found in-person meetings challenging to attend:

> *And it’s just amazing like, there’s so much available. So I can do one any time of any day and it’s really accessible and with having schizophrenia you know, and anxiety disorder, sometimes it’s quite difficult for me to make it to the physical meetings* (client 10, M, 41-45)

Service providers reported how the use of remote services, including assessments and online psychosocial sessions, were also beneficial in terms of treatment engagement and outcomes. It was suggested that the increased accessibility that remote service provision provided reduced occurrences of missed appointments, which were previously met with punitive responses:

> *It gave them ownership and flexibility of their own time and their own treatment. I’d say that previously we were quite strict on, you know you have to attend this appointment every four weeks and you have to do this. Whereas actually I think for some people, if when it went over the phone a lot of them really appreciated it* (team leader, F, 26-30)

One provider also suggested that the transition to remote and digital services, and the benefits this afforded, resulted in increased contact rates during the pandemic:

> *I would guess if we were to look at contact rates, they were probably a little bit better during the pandemic than they probably would have been beforehand just because people could pick up a phone a lot easier than they can get into the service* (consultant psychiatrist, M, 36-40)

In contrast, others welcomed the return to in-person services, particularly as several challenges and issues with remote service provision were reported. For instance, most clients described connectivity issues, citing a lack of phone or internet as significant barriers to engagement:

> *But a lot of people don’t have that access, your normal heroin user who injects fucking five bags of heroin a day and 10 pieces of crack a day, he’s not going to have an iPad where he can go on a Zoom meeting* (client 10, M, 41-45).

Despite one of the services reporting the handout of mobile phones to address this issue, providers reported limitations for treatment quality. Often this was related to an inability to adequately assess the physical health of clients via other sensory cues (*I felt you lose a lot from not having that contact with people. There’s information that can get missed, people probably wouldn’t be as open over the phone…you get limited information through phone contact as opposed to face to face*, consultant psychiatrist, M, 36-40). Providers therefore felt that a lack of face-to-face contact made it easier for clients to provide false or misleading information regarding their substance use:

> *It’s a lot harder to assess people over the phone. Obviously they will tell us what they want to tell us, which is easier over the phone. What I’ve noticed, once I was able to see people face-to-face, the presented in person was very different from the way they presented over the phone*., *“Oh yes, I’m not too bad”, and they come into the office and they’re actually yellow* (recovery coordinator, F, 51-55)

Some clients were keen to return to in-person group sessions that provided a sense of routine, structure and companionship. For example, one client described how the connections and camaraderie she shared with peers in previous face-to-face group sessions were not achievable through remote online group sessions:

> *I used to go to the sex work drop-in and see other sex workers. And I loved it. We used to sit and chat and slag off clients, blow up condoms, have a vent. Slag off our mutual clients, and moan about our job. And see support services, grab condoms, something to eat, have a natter*.*…because when you’re at the drop-in, it’s easier, we can talk. But I can’t ring up my mate and be like just started chatting about. I don’t know who she’s with. I don’t know if she’s able to talk about it. She might be with her partner who doesn’t know. So, that made it impossible. And it’s made it a really lonely experience* (client 3, F, 21-25)

As services weighed up whether to continue with forms of remote service provision, there was some suggestion that the quality of this treatment hinged on service provider comfort in delivering services via this method. Indeed, some providers described variability in responses among staff to providing remote provision: of those familiar and comfortable with responding to clients in this way (*some people were tech savvy and very keen to do it and very proactive and set things up*) and those less so (*others had to be really pushed to do it because it was something new and something they weren’t comfortable with*, Services Manager, M, 41-45). Some service providers also described feeling inadequately equipped to deal with often challenging and complex issues experienced by clients over the phone, which, at times, led to feelings of powerlessness:

> *People are more and more depressed, people are phoning up saying, “I’m going to kill myself”, we’re having to deal with that. We’re not trained to deal with that. But people just phoning up in situations where I’m actually powerless to help them and, “Here’s the number for the Samaritans and I’ll refer you to be put on the waiting list for the service you need”* (recovery coordinator, F, 51-55)

## Discussion

Our findings are valuable, as to date, little is known about the ongoing impact of the pandemic on the health, wellbeing and daily lives of PWID. Furthermore, the experiences of service providers responding to the challenges faced by PWID are mainly absent, particularly in the UK where – to our knowledge – qualitative research has focused exclusively on the perspectives of clients (Kesten et al., 2021). Therefore, this study provides new insights into the social and psychological impact of the COVID-19 pandemic on PWID and their experiences of service adaptations in this context.

Our findings suggest PWID remain fearful of COVID-19, yet broader socio-structural inequalities, including accommodation conditions and economic hardship, are limiting the ability to comply with public health measures. This confirms earlier quantitative findings that socioeconomically vulnerable populations and PWID have fewer possibilities to adhere to guidelines (Beale et al., 2021; Genberg et al., 2021). Our data adds qualitative insights into what these limited possibilities may be and how they are continuing to affect this population. For instance, the risk of financial and material hardship and the daily burden of keeping well compelled some to disregard social distancing despite risks of COVID-19 transmission, as evidenced by a continued engagement in street-based sex work among some clients. As reported elsewhere (e.g. Kesten et al., 2021), accommodation settings and living arrangements were also unconducive to social distancing, which we also found elevated distress by limiting the contexts in which physical distancing could be practised.

Feelings of vulnerability to infection have led to poorer mental health among other population groups: those with long-term health conditions, for example, reported fear and anxiety related to the consequences of COVID-19 infection (Fisher, Roberts, McKinlay, Fancourt, & Burton, 2021). Our findings suggest a similar, ongoing occurrence among PWID. Any additional burden on mental health among this population is particularly concerning given PWID already experience a high prevalence of mental health symptoms and barriers to accessing mental health services (Genberg et al., 2019; Priester et al., 2016) as well as it being a risk factor for ongoing substance use (Pilowsky, Wu, Burchett, Blazer, & Ling, 2011). Hence, whilst these findings highlight the importance of accessible mental health support in pandemic response measures - including access to low-threshold, co-located psychological services - there is a need for additional support attending to the social-structural vulnerabilities that shape COVID-19 related harms, including appropriate housing, shelter, food and economic support.

Our findings provide further evidence of how COVID-19 has altered a number of indices of drug-related harm among PWID, including physical (e.g. changes in drug use patterns and characteristics) and socio-economic (e.g. riskier sex work engagement and income generation) harms. Volatility in the global heroin supply and reduced availability and quality of substances at local level have been observed during the pandemic (Bennett et al., 2021). Difficulties in sourcing drugs is particularly problematic as there is evidence that PWID may substitute or transition to more readily available but unfamiliar substances in attempts to abate withdrawal symptoms (Day et al., 2003; Harris, Forseth, & Rhodes, 2015; May, Holloway, Buhociu, & Hills, 2020). Our findings, along with earlier data from the pandemic (Croxford et al., 2021; Kesten et al., 2021; Morin et al., 2021), suggest that similar patterns may have occurred due to accessibility issues. However, our data adds further insight in that some changes to drug use appeared to be temporary, with clients reverting to the sole use of heroin as availability returned. In contrast, there was some evidence to suggest the development of increased tolerance and dependence on substances initiated in response to shortages during the earlier stages of the pandemic, including crack cocaine, benzodiazepines and pregabalin. Although we did not identify any occurrences of significant harm, the observed shifts to increased depressant polydrug use is a concern given the potential for harm (including increased vulnerability to overdose, BBV infection (Harris et al., 2015; Horyniak et al., 2015)), particularly if used alongside opioids (Macleod et al., 2019; McAuley, Matheson, & Robertson, 2022). Switches to crack cocaine use during this period are also problematic given users’ proclivity to share pipes, increasing COVID-19 transmission risks (Harris, 2020). In this context, increased take-home naloxone distribution, removing barriers to treatment, public health messaging, and access to blood-borne virus testing are essential to minimize various drug-related harms associated with limited availability, including withdrawal. The changes to crack-cocaine reported by participants in our study and the increasing availability and prevalence of its use during the pandemic more broadly (EMCDDA., 2021) also necessitate harm reduction measures to help reduce COVID-19 infection and respiratory-based health harms among this population of users (Harris, 2020).

Opportunities for informal and illegal income generation were also reportedly reduced by the pandemic and found to impact the wellbeing of PWID. Interview narratives suggested those most impacted were women reliant on survival sex work due to their increased economic precarity during this period and limited access to social protection (Platt et al., 2020). There is an extensive body of research drawing attention to how economic vulnerability renders this population more susceptible to health-related harms, particularly if drug use is part of this dynamic (Ogden et al., 2021). This includes reduced possibilities to negotiate prices and screen clients, which may contribute to unprotected sex and gender-based violence, both of which are risk factors for HIV (Strathdee et al., 2015).

Our findings suggest the pandemic has compounded many of these issues and heightened the possibility of these harms occurring in this context. For instance, a reduced demand for street-based sex work and increased competition from online sexual services in the immediate period following the relaxation of social distancing measures severely limited some sex-workers agency over transactions, including capacity for safer sex practices, client screening and price negotiation. These hazards played out alongside other risks, with some women obliged to enter into relationships with male partners for economic support. These women were especially vulnerable to exploitation given how gender inequitable power relations often play out in drug-using partnerships, notably through the control of resources generated through sex work (Bourgois, Prince, & Moss, 2004). Hence, there is an urgent need for financial support and protection for those with no option but to continue sex working during this period (Platt et al., 2020).

COVID-19 has created a context in which innovative service responses and regulatory changes can be implemented and examined, thereby providing preliminary insight into the feasibility of future service adaptations. A shift to weekly or fortnightly OST pickups during the earlier stages of the pandemic were well received and afforded a number of benefits to clients, including stigma reduction, increased day-to-day freedom, and greater control over daily dosage, as reported elsewhere (Krawczyk, Fawole, Yang, & Tofighi, 2021; Nordeck et al., 2021). Building on these earlier findings, our data provides insights into how clients and providers perceived changes back to daily supervised consumption following a relaxation in social distancing rules. Most clients expressed displeasure at this reversal given the aforementioned benefits and a perception that they had ‘proved’ themselves capable of managing take-home doses. This is largely in contrast to a continued scepticism and caution among providers toward any sustained relaxation of regulations, as similarly reported elsewhere (Hunter et al., 2021; Krawczyk et al., 2021). Whilst these contradictions reflect long-standing debate between client/provider preferences regarding the optimal delivery of OST (Frank, 2021), services may wish to take stock of the benefits afforded from these changes during this period (e.g. Figgatt et al., 2021; Frank et al., 2021; Kesten et al., 2021). This is especially important given the daily burden of supervised consumption was often cited as one reason behind low retention rates in OST prior to the pandemic (Frank, 2021; Nolan et al., 2015). Further research is required to enable best practice in balancing the potential risks and benefits of relaxed regulations, including objective patient outcome data to determine whether flexibilities have improved or worsened treatment outcomes, medication diversion and overdose.

Whilst remote service provision, including the use of telephonic methods, were implemented rapidly to increase service access, most services reported retaining some elements of remote operating at the time of data collection and envisioned their continued use in the future. Although providers and some clients reported benefits of virtual forms of treatment, including increased access and the ability to reach clients unwilling to engage with face-to-face psychosocial services, our findings also highlighted several disruptions to care due to the ongoing use of such methods. These were mostly related to difficulties assessing client health and wellbeing virtually and perceived reductions in levels of support provided. This is in line with earlier research reporting a lack of provider comfort and willingness in using such methods (Aronowitz et al., 2021; Goldsamt, Rosenblum, Appel, Paris, & Nazia, 2021), and disrupted support routines among clients (Kesten et al., 2021). PWID also face disparities in accessing telehealth services, and although innovations – including onsite, private rooms with sanitized telephones (Quiñones et al., 2021) and the distribution of donated mobile phones (Komaromy et al., 2021) – have proved successful in offsetting some of these issues, there remains a need to ensure access to telehealth services is more evenly distributed among PWID. Similarly, providers must be equipped and comfortable with using telehealth methods as their use becomes increasingly integrated in treatment frameworks and routine health care (Aronowitz et al., 2021). The development of an improved telehealth infrastructure, including dedicated and tailored telehealth training curricula for health and social care workers, has been recommended (Fisk, Livingstone, & Pit, 2020; Thomas et al., 2020), and may have similar utility for those working in drug services. Our findings also suggest the need for further research into the effectiveness of remote service provision on client engagement and treatment outcomes.

## Limitations

This study is not without limitations. First, the sampling strategy may be biased toward those participants willing or able to participate. It is possible the views expressed in this study differ from those unwilling or unable to participate and may contribute to the under-reporting of certain experiences (e.g. overdose). Second, client and provider interviews were conducted over several months and may therefore reflect the impact of time-specific events or experiences, including lockdown measures or service alterations. For example, interviews were conducted at a time when COVID-19 legal restrictions were lifted in UK, including the removal of social distancing and social contact limits and the reopening of businesses (Institute for Government Analysis, 2021; The Scottish Parliament, 2022). The timing of interviews therefore require consideration when interpreting the findings. Nevertheless, participants were able to recount both current and retrospective experiences during periods when more restrictive social distancing measures were in place (e.g. stay at home orders). Third, whilst the study includes the experiences of providers and clients from various regions of England and Scotland, regional differences in service provision, drug markets and lockdown measures may mean perspectives and experiences vary in ways that are not fully captured in this research. Finally, despite efforts to recruit a geographically diverse sample, the majority of service users were recruited from a single service. This is perhaps reflective of the challenges recruiting participants for qualitative studies during the pandemic, particularly from organisations or services that have been under significant pressure to adapt and manage services since its onset. In attempts to make data collection as non-burdensome and straightforward for organisations as possible, a convenience sampling method was therefore adopted, resulting in the collection of data from participants most accessible at the time of the study. However, as there were also difficulties conducting remote interviews with the study population during this period (e.g. limited phone ownership, housing instability, see also Parkin et al. (2021)), it was necessary for the research team to find ways to facilitate the full participation of this population. This entailed conducting some face-to-face interviews at services accessible (geographically and logistically) to the research team. While the geographic makeup of service users in this study is therefore somewhat distorted, it does include the insights and experiences of those who would not have been able to participate in the study if it were conducted entirely via remote methods. The inclusion of service providers in our study meant we could also involve geographically diverse perspectives, given that no recruitment issues were experienced with this group.

Despite these limitations, the paper is the first known study in the UK to interview both drug service providers and PWID about their ongoing experiences of the COVID-19 pandemic. Understanding the experiences of the latter is particularly important given their increased marginalisation and vulnerability during this period. Thus, the study provides a voice to a seldom heard group through in-depth, semi-structured interviews, which can provide important insights for future policy directions and service provision.

## Conclusion

This paper provides new insights into the ongoing impact of the pandemic on the mental health, drug-related harms and behaviours of PWID as well as adaptations to treatments and services. Whilst our findings emphasize the importance of accessible support measures attending to the immediate priorities of PWID during this period (including access to low-threshold, co-located psychological services, naloxone, NSP provision, low-threshold OST programmes) there is a need for additional support addressing the social-structural vulnerabilities that disproportionality affect PWID. This includes the provision of appropriate housing, shelter, food and economic support to ensure pre-existing disparities and harms are not exacerbated further by the conditions of the pandemic. Drug service adaptations initiated in response to the pandemic also require further attention to ensure future treatment is acceptable and ultimately responsive to the needs of PWID. In this context, it is important that any innovations in treatment are informed by the knowledge and expertise of PWID themselves, given the benefits afforded by some service adaptations (e.g. relaxation of OST). However, service deliverers may require further support (e.g. tailored training curricula) to increase acceptance of any sustained policy and service delivery adaptations prompted by COVID-19.

## Data Availability

The datasets presented in this article are not readily available because they contain information that could compromise the privacy of research participants. Requests to access the datasets should be directed to

## Author Contributions

AB and DF contributed to the conception and design of the study. TM was responsible for data collection and wrote the first draft of the manuscript. TM and JD performed the formal data analysis. AB, DF, and JD assisted with review and editing. All authors contributed to manuscript revision, read, and approved the submitted version.

## Acknowledgements

The authors would also like to thank the organisations that assisted in the recruitment of participants for this study, as well as those people who gave up their time to take part and contribute to the study. The authors would also like to thank Dr Alison McKinlay and Dr Katey Warran for their assistance during the recruitment and analysis stages of this paper.

## Footnotes

1 Clinical guidance for commissioners and providers of drug and alcohol services regarding the safe delivery of OST during the pandemic, including possibilities of risk-assessed, relaxed pick-up and supervision requirements, was withdrawn on 19^th^ July 2021 (Department of Health & Social Care, 2021).

## Notes

### Competing Interest Statement

The authors have declared no competing interest.

### Funding Statement

The COVID-19 Social Study was funded by the Nuffield Foundation (WEL/ FR-000022583), but
the views expressed here are those of the authors. The study was also supported by the MARCH
Mental Health Network funded by the CrossDisciplinary Mental Health Network Plus initiative
supported by UK Research and Innovation (ES/S002588/1) and by the Wellcome Trust
(221400/Z/20/Z). DF was funded by the Wellcome Trust (205407/Z/16/Z).

### Author Declarations

The study was reviewed and approved by the University College London Ethics Committee (Project ID 6357/002)

