## Appendix 1 for "A qualitative study exploring the impact of the COVID-19 pandemic on People Who Inject Drugs (PWID) and drug service provision in the UK: PWID and service provider perspectives"

Appendix B: Covid-19 restrictions during 2021 in England and Scotland (up until completion of data collection period (September 2021))*

| **Date** | **Covid-19 restrictions** |
| --- | --- |
| 5/6^th^ January 2021 | - Third national lockdown begins in England and Scotland. |
| 8^th^ March 2021 | - Schools in England reopen for primary and secondary school students. |
| 29^th^ March | - ‘Stay at home’ orders end in England, although people encouraged to remain local. - Outdoor gatherings with either six people or two households permitted in England. Outdoor sports facilities also reopen. |
| 2^nd^ April 2021 | - ‘Stay at home’ orders end in Scotland, allowing non-essential journeys in local authority areas. |
| 5^th^ April 2021 | - Opening of non-essential retail, including hairdressers, libraries, beauty salons and museums in Scotland |
| 12^th^ April 2021 | - Non-essential retail, hairdressers, public buildings (e.g. libraries and museums) reopen in England. - Outdoor venues, including pubs, restaurants and themes parks, and some indoor leisure settings (e.g. gyms) also reopened. |
| 26^th^ April 2021 | - Easing of restrictions across retail and hospitality in Scotland, allowing cafés, pubs and restaurants to resume full outdoor service and serve food indoors without alcohol until 8pm - Non-essential travel between Scotland and other Home Nations also permitted. |
| 17^th^ May 2021 | - Two households or ‘rule of six’ permitted to gather indoors for social gatherings in England. Indoor economy, including pubs, restaurants and cinemas, also reopened. - Up to six people from three households permitted to meet in each other’s homes or gardens with physical distancing in Scotland (with the exception of Moray and Glasgow) - Opening of indoor economy, including pubs, restaurants and cinemas in England and Scotland. Pubs and restaurants permitted to serve alcohol until 10:30pm in Scotland |
| 19^th^ July 2021 | - Removal of majority of legal restrictions on social contact in England and Scotland, and sectors of the economy that were previously closed now reopened (e.g. nightclubs). Hospitality venues in Scotland permitted to open until midnight |
| 9^th^ August 2021 | - Remaining COVID restrictions (legal requirements for physical distancing and limits on gatherings) lifted in Scotland, although compulsory mask wearing in some locations remain. Nightclubs also permitted to reopen |

*(Institute for Government Analysis, 2021; The Scottish Parliament, 2022)

Institute for Government Analysis. (2021). Timeline of UK government coronavirus lockdowns and measures, March 2020 to December 2021 Retrieved from <https://www.instituteforgovernment.org.uk/sites/default/files/timeline-coronavirus-lockdown-december-2021.pdf>

The Scottish Parliament. (2022). Timeline of Coronavirus (COVID-19) in Scotland. Retrieved from <https://spice-spotlight.scot/2022/03/18/timeline-of-coronavirus-covid-19-in-scotland/>
