## Appendix 2 for "A qualitative study exploring the impact of the COVID-19 pandemic on People Who Inject Drugs (PWID) and drug service provision in the UK: PWID and service provider perspectives"

**Interview topic guide: Substance misuse groups**

**Ask to describe ‘normal life’**

- Employed? Type of job, hours etc,
- Living situation
  - Who you normally live with, does this change, separated/ extended family?
  - Has this accommodation changed recently?
  - Do you feel secure in your accommodation? Do you have options to go elsewhere if needed?
  - Have your living arrangements changed during the pandemic?
- Education/study?
- Full time parent or carer?
- Use of any community services?
- If they have any health conditions they mentioned (what condition, when diagnosed, if on or whether they have had complete treatment)
  - What was current treatment plan? How was it being managed? What was usual routine for appointments/follow-up?
- Whether you would usually have done any type(s) of regular exercise (whatever they perceive as exercise including walking/gardening)
- Are you currently registered with any support services? (third sector etc).

SUBSTANCE USE

- **Please describe your current substance use?**
  - Substance(s)? or Alcohol? (preferred drink? – beers/spirits etc?)
  - Frequency and quantity? Cost? How much/often?
  - Period of problem substance use? (months/years)

If opioid user,,,

- Method of administration – intravenous/intranasal/ smoke?
- Any harm reduction practices? (e.g. with peers, no syringe sharing?)
- **Was there any impact of the pandemic on your substance use?**
  - Getting hold of substances (ability to purchase drugs/alcohol etc).
  - Substance(s)? Any change in patterns of substance use? (e.g. switch to alternative substances or different alcoholic beverages? If so, why?)
  - Any increase in frequency/quantity/or cost?
- If opioid user…
  - Changes in methods of administration? (if so, why? Availability of equipment?)
  - Changes in harm reduction practices? (e.g. with peers/without, sharing syringes?)
  - Any changes in other parts of life that link with substance use? (e.g housing/relationships)
- **Are you receiving any treatment for your substance use?**
  - Substitute medication (e.g. methadone/Subutex?)
    - Detox/maintenance? Dosage? Length of time in treatment?
  - Any pharmacological treatment medications to ease alcohol withdrawal if detoxing? (e.g chlordiazepoxide).

Home/clinical detox?

- Any other support? (e.g. psychosocial)
- **Was there any impact of the pandemic on your ability to receive/access treatment/support? (e.g OST/NSP)**
  - Any changes in the support/treatment you receive? (e.g. non-daily pickup?
  - Changes in operational practices/modes of treatment (e.g. communication methods?)
  - Engagement/relationship with services?
  - Is there anything that support services could have done better?

UNDERSTANDING AND ADHERENCE TO GUIDELINES

**At the moment, are you self-isolating (how long for, reasons for this) a key worker, working but not a key worker, social distancing/ ‘staying at home’**

- Please describe what this is for you and your family/ household?
  - i.e. are you self-isolating with neighbours helping to get groceries, or going out for these?
  - Or self-isolating with no outside exercise? Etc – if self-isolating without outside exercise how are you feeling about this? Do you have a garden or outdoor space?
- **What do you understand by the ‘social distancing’ advice that is being given – what does it mean to you?** Have you been…
  - Avoiding crowds
  - Keeping personal distance from others
  - Isolating
  - Avoiding close contact greetings
  - Socialising/going out only with those in your household
  - <if exercising outside> are you finding places to go where you can keep your distance from others?

**Have you been able to stick to the social distancing advice that has been given to your group? Please tell us about why/ why not?**

**[COM-B prompts can be used here, to include:]**

- Have your experiences (e.g. substance use (**OR** domestic abuse/financial hardship/ homelessness)) had any impact on being able to follow social distancing guidelines? How?
- Any existing physical or mental health problems
- Group membership/ applicability
- Beliefs about consequences/ health beliefs
- Consequences for others/ self
- Needing to work/ living arrangements, whether others are self-distancing in the same house/area
- Work/ Caring responsibilities, providing emotional support
- Peer pressure to socialise
- Government rules/punishments <prompt to ask how they feel about the Government recommendations that are relevant to exercise for them>,
- Feelings about losing normal life
- Change of routine/ habits
- Any impact on your ability to access support?

HEALTH CONDITIONS

**How has Covid-19 had an impact on the [health condition] AND/OR specific health needs (e.g. compromised immunity, mental health).**

Prompts include:

- What has been the impact on any normal appointments? (cancelled, delayed, unable to speak to appropriate healthcare professional, changed to different method/location of appointment e.g. online/telephone)
- What has been the impact on any treatment? (cancelled, delayed, changed from usual treatment plan, unable to get medication/prescriptions)
- Have you experienced an impact on any symptoms/side effects?
- How have you felt about [any mentioned changes/impact above]?

**SOCIAL LIFE**

**How would you describe your social life before the Covid-19 pandemic?**

- How would you describe your social network – for example size, types of people, types of relationships, do they live with you, nearby or further away, how often do you see each other, how well do you know each other? How do you interact, face to face, online or social media?
- Social activities?
- Could you describe any community services/participation or volunteering participation?
- Could you describe the social support you have? (such as emotional support, advice and information, someone to help you with money or milk/bread/essentials, community services)
- Can you tell us about any ways your social networks/ friendship groups influence you, such as peer pressure, or encouraging you to get involved in things? Do you compare your life to theirs?
- Social engagement (social roles, bonding, attachment)

**How would you describe your social life now that social distancing measures have been brought in because of Covid-19? Please tell us about this**

Prompts include:

- How would you describe your social network – for example size, types of people, types of relationships, do they live with you, nearby or further away, how often do you see each other, how well do you know each other? How do you interact, face to face, online or social media?
- Social activities?
- Could you describe any community services/participation or volunteering participation?
- Could you describe the social support you have? (such as emotional support, advice and information, someone to help you with money or milk/bread/essentials, getting medication/access to healthcare, community services)
- Can you tell us about any ways your social networks/ friendship groups influence you, such as peer pressure, or encouraging you to get involved in things? Do you compare your life to theirs?
- Social engagement (social roles, bonding, attachment)
- How has your experience of substance misuse (**OR** domestic abuse/financial hardship/homelessness) during the pandemic impacted your social life/networks?

MENTAL HEALTH

**How do you feel about the changes that have been brought about by Covid-19?**

**Have they had any impact on your mental health or wellbeing? Please tell us about these**

- What are the things most bothering you at the moment?
- Have you experienced any impact on positive emotions? (prompts: how deeply you can engage with what you are doing, sense of meaning/ purpose, relationships with others, how well you are managing and feelings of control over your situation?)
- Has there been any impact on your sense of identity?
- Have you experienced any negative psychological feelings? (prompts: such as shame, guilt, lack of pleasure, anxiety, worry)
- Please tell us about any physical symptoms due to being stressed or anxious? (prompts: fatigue, sleep problems, pain, illness symptoms, palpitations)
- Has your experience of using substances **OR** domestic abuse/financial hardship/homelessness during the pandemic affected your mental health and wellbeing? If so, how?

**Have you been doing/ planning anything to help with this?**

- Connecting with family or friends/ work colleagues online?
- Online groups?
- Hobbies/ Reading
- Exercise at home <ask about what they have been doing and if there are specific resources they have found useful to exercise>
- Volunteering
- Other engagement

**Why are you doing/ not doing these things?**

- Helpful/ not helpful – please tell us why
- Enjoyable
- Good for mental health/ wellbeing
- Can’t get online, not connected, not comfortable, affordability, confidence in using/ skills
- Skills in using the internet/ communication software
- Living arrangements/ Work/ caring demands
- Peer support/ pressure
- Difficulties/ restriction in physical environment

PROSPECTION

**Has the pandemic meant that you have any worries for the future?**

**How are these different from the worries you had before?**

- Sense of control/ powerlessness
- Severity of worries / perspective

**Will this change the way you live your life in future?**

- The way you connect with others
- How you look after yourself
- How you support others
- How you work?
- How you exercise?

Has this changed any of your priorities for the future?
