## Appendix 3 for "A qualitative study exploring the impact of the COVID-19 pandemic on People Who Inject Drugs (PWID) and drug service provision in the UK: PWID and service provider perspectives"

**Welfare Providers (substance use/homeless/domestic abuse/employment or financial support)**

**INTRODUCTION**

**Ask to describe work/type of service**

- Type of service?
- Groups/clients that you work with?
- What is your role?
- What services/practices/initiatives do you offer?

**IMPACT ON CLIENT GROUP**

**What are the key issues facing your client group (e.g. homeless, substance use, intimate partner violence) during the pandemic?**

- Have your clients been able to understand and adhere to the social distancing guidelines? (If yes - why/what has helped?/If no why not?)
- Have their experiences (e.g. homelessness/substance misuse/intimate partner violence) had any impact on being able to follow social distancing guidelines?
- Any existing physical or mental health problems
- Beliefs about consequences/ health beliefs
- Consequences for others/ self
- Needing to work/living arrangements, whether others are self-distancing in the same house/area
- Work/ Caring responsibilities, providing emotional support
- Peer pressure to socialise
- Managing to follow advice re facemasks indoors/ handwashing etc
- Government rules/punishments/messaging
- Feelings about losing normal life
- Change of routine/ habits
- What has been the impact of the pandemic on your clients being able to access services?
  - What has been the impact on any normal appointments for your group? (cancelled, delayed, unable to speak to appropriate support workers or health professionals)
  - Has there been an impact on any treatment? (cancelled, delayed, changed from usual treatment plan, unable to get medication/prescriptions)
- Have you noticed any impact on their physical/mental health? Could you describe some examples?
  - increased symptoms/side effects
  - Increase or change in presentation of conditions?
  - Impact on client ability to manage physical/mental health?
- Have there been any changes to the living arrangements of your clients? Could you describe some examples?
- Have your clients had any difficulties accessing daily provisions (including sanitation, hygiene products, food)? Could you describe some examples? Has anything helped them to access these provisions?
- **Substance use (only if noted or in substance use groups)**
  - Substance(s)? Have you noticed any changes in patterns of substance use? (e.g. switch to alternative substances/beverages)? If so, why?)
  - Any increase in frequency/quantity/cost? Methods of purchasing/obtaining?
- If working with opioid users…
  - Changes in methods of administration? (if so, why? Availability of equipment?)
  - Changes in harm reduction practices? (e.g. with peers/without, sharing syringes?)
  - Any changes in other parts of clients life that link with substance use? (e.g housing/relationships)

**CHALLENGES TO OPERATIONAL PRACTICES/SERVICE DELIVERY**

**What are the key challenges in delivering your service during the pandemic?**

- Ability to follow rules around social distancing and maintain delivery of service?
- Changes in services due to organisational or gov guidelines?
- Safety of staff/service users?
- Maintaining contact/relationships with service users?
- Impact on/emotional wellbeing of staff?
- Use of PPE and other measures to protect staff from the virus?

**Has your service made any adaptations in response to the challenges encountered during the pandemic?**

- Any changes to current practices during lockdown to deal with issues?
- New initiatives? (e.g. digital comms, buvidal, temp accommodation)
- Different methods of communication?
- Location of appointment? (e.g. online/telephone)
- Any benefits to come out of changes from the pandemic?

**PROSEPECTION**

**Has the pandemic meant that you have any worries for the future (for your client group)?**

- Are there likely to be any lasting impacts/consequences of the pandemic on your clients?
- Are there likely to be any lasting impacts/consequences of the pandemic on service provision?

**How are these different from the worries you had before the pandemic?**

**Will this change the way you deliver your service in the future?**

- The way you connect with service users?
- How you support service users?
- How you operate daily
- The services/initiatives/practices that you deliver?
- How you support staff?

**Has this changed your priorities as a service for the future?**
